# Adoption of the World Health Organization’s best practices in clinical trial registration and reporting among top public and philanthropic funders of medical research in the United States

**DOI:** 10.1101/2023.04.03.23288059

**Authors:** Elise Gamertsfelder, Netzahualpilli Delgado Figueroa, Sarai Keestra, Alan Silva, Ronak Borana, Maximilian Siebert, Till Bruckner

## Abstract

**Background/Aims:** Clinical trial funders in the United States have the opportunity to promote transparency, reduce research waste, and prevent publication bias by adopting policies that require grantees to appropriately preregister trials and report their results, as well as monitor trialists’ registration and reporting compliance. This paper has three aims: a) to assess to what extent the clinical trial policies and monitoring systems of the 14 largest public and philanthropic medical research funders in the United States meet global best practice benchmarks as stipulated by the WHO Joint Statement;[1] b) to assess whether public or philanthropic funders have adopted more WHO Joint Statement elements on average; and c) to assess whether and how funders’ policies refer to CONSORT standards for clinical trial outcome reporting in academic journals.

**Methods:** The funders were assessed using an 11-item scoring tool based on WHO Joint Statement benchmarks. These 11 items fell into four categories: trial registration, academic publication, monitoring, and sanctions. An additional item captured whether and how funders referred to CONSORT within their trial policies. Each funder was independently assessed by 2-3 researchers. Funders were contacted to flag possible errors and omissions. Ambiguous or difficult to score items were settled by an independent adjudicator.

**Findings:** Our cross-sectional study of the 14 largest public and philanthropic funders in the US finds that on average, funders have only implemented 4.1/11 (37%) of World Health Organization best practices in clinical trial transparency. The most frequently adopted requirement was open access publishing (14/14 funders), and the least frequently adopted were (1) requiring trial ID to appear in all publications (2/14 funders, 14%) and (2) making compliance reports public (2/14 funders, 14%). Public funders, on average, adopted more policy elements (5.3/11 items, 48%) than philanthropic funders (2.8/11, 25%). Only one funder’s policy documents mentioned the CONSORT statement.

**Conclusions:** There is significant variation between the number of best practice policy items adopted by medical research funders in the United States. Many funders fell significantly short of WHO Joint Statement benchmarks. Each funder could benefit from policy revision and strengthening.

## Background

Prospective clinical trial registration and timely, public disclosure of trial results are of utmost scientific, ethical, and financial importance.[1] Prescribers, patients, and public health officials rely on complete and accurate trial information for the treatment of disease. Failure to register clinical trials and report results can lead to needless duplication and research waste.[2] One estimate suggests that up to 85% of the money spent on medical research globally is wasted, half of which is due to non-reporting of results.[3]

Most of this waste is avoidable. Improvements in trial design, strengthened regulatory requirements, and increased oversight can help curb waste. Because strengthening regulatory power at a national level is difficult and legislative change happens slowly, individual funders of medical research are in a unique position to improve research policy at the ground level. By requiring their grantees to meet specific criteria as a condition of funding, as well as by monitoring grantee compliance, funders can reduce waste in clinical research even if national regulators fail.

In addition to waste, poor clinical trial practices can lead to publication bias. Failure to publish both positive and negative outcomes of trials affects the availability of evidence for prescribers and the public.[4] Negative trial results go unreported more frequently than positive results,[5] leading to a distortion of evidence that overstates the efficacy of new drugs, medical devices, and technologies, while downplaying their harms.[6]

Appropriate trial registration and reporting is also an ethical imperative: publication bias undermines regulatory decision-making, inhibits the development of clinical guidelines, and interferes with health technology assessment.[6] For this reason, at the 2013 UN General Assembly, the World Medical Association expanded the Declaration of Helsinki (a set of ethical principles guiding human subjects research) to include a new imperative: reporting negative and inconclusive research findings as well as positive ones.[7]

Expanding on the Declaration of Helsinki, the World Health Organization (WHO) released a statement in 2017 outlining global best practices for clinical trial registration and reporting. The “Joint statement on public disclosure of results from clinical trials” (hereafter “WHO Joint Statement”) has been signed by 23 major medical research funders, each pledging to reduce waste, curb publication bias, and advance scientific progress through strengthened clinical trial policies.[1] Just one of the 14 funders in this cohort (the Bill and Melinda Gates Foundation) is a signatory. The WHO Joint Statement encourages funders of clinical trials to ensure their grantees preregister their trials and post results on the same registry within 12 months of trial completion. It also asks funders to monitor registration and reporting compliance and to make monitoring reports public. The 2022 World Health Assembly adopted a global resolution to bolster clinical trial quality and transparency, referencing the WHO Joint Statement standards within the resolution.[8]

The WHO Joint Statement thus provides a global benchmark for registration and results reporting in clinical research. Clinical research standards vary significantly between countries - the US’ National Institute of Allergy and Infectious Disease runs a helpful website that compares regulations across 20+ countries[9] - but regardless of location, all human subject research should be held to the same high standards. The WHO Joint Statement has specific policy elements that can be universally applied and enforced.

In the United States, registration of a limited subset of clinical trials has been a legal requirement since Congress’ passage of the 1997 Food and Drug Administration (FDA) Modernization Act.[10] This resulted in the creation of ClinicalTrials.gov, the largest database of clinical studies in the world, maintained by the National Library of Medicine, a subsidiary of the National Institutes of Health (NIH).[11] Clinical trial submission requirements were expanded in 2007 with the passage of the FDA Amendments Act (FDAAA), requiring both registration and results for applicable trials to be submitted to the ClinicalTrials.gov database. Under FDAAA, trials must be registered within 21 days of initiation and results must be posted within 12 months of study completion or termination.

The FDAAA also introduced financial penalties for results submission noncompliance, up to a maximum of US$13,000 per day after receiving a Notice of Noncompliance.[12] To date, the FDA has only ever threatened four noncompliers with a fine,[13] but could have imposed penalties totaling over US$34 billion since the FDAAA became enforceable in January 2017.[14]

All applicable clinical trials (ACT) in the US, regardless of funding, must abide by the FDAAA regulations. Even so, the ACT criteria covers only a small minority of interventional trials.[15] Gaps in the legal framework and regulatory enforcement provide a strong rationale for funders to insist that their grantees register and report all interventional trials. As former NIH Director Francis Collins put it, “It’s hard to herd cats, but you can… take their food away.”[16]

Federally funded studies are subject to additional requirements for data collection and dissemination.[9] Grantees funded by any agency of the Department of Health and Human Services, including the NIH, FDA, and Agency for Healthcare Research and Quality (AHRQ), must register and submit ACT results as a condition for continued and future funding.[17] Complementary to this, the NIH issued a dissemination policy that covers all NIH-funded trials, not just ACTs.[18] However, as found in this and several other studies,[19–21] legal requirements, ethical considerations, and reality do not always coincide. Notably, federally funded studies were found to be significantly less likely to adhere to FDAAA mandates than industry sponsored trials.[22] NIH-funded studies are reported within 12 months of study completion just 8% of the time, while other federally funded studies have a 12-month results reporting rate of 5.7%.[23]

## Rationale

This study builds on prior research that reveals significant gaps between WHO Joint Statement benchmarks and the policies of major medical research funders in the US and Europe.[19–21, 24] This study differs from prior studies of US funders by specifically assessing policy elements contained in the WHO Joint Statement, as well as including funders not previously assessed.[20]

## Objectives

The primary objective of this study is to assess the extent to which the clinical trial policies and monitoring systems of the 14 largest public and philanthropic medical research funders in the United States meet global best practice benchmarks as outlined in the WHO Joint Statement. The secondary objectives are to assess a) whether, on average, public or philanthropic funders in the US have adopted more policy items and b) whether and how funders’ policies refer to the Consolidated Standards of Reporting Trials (CONSORT) standards for clinical trial reporting in journals.[25] Though CONSORT is not mentioned in the WHO Joint Statement, it has been endorsed by nearly 600 journals and organizations worldwide.[26]

## Methods

We closely followed the Bruckner et al. (2022) protocol (which assessed the clinical trial policies of European funders) and retained the original assessment tool and rating guide. The assessment tool is an 11-item based on WHO Joint Statement benchmarks. The 11 items fell into four categories: trial registration, academic publication, monitoring, and sanctions. An additional item captured whether and how funders referred to CONSORT within their trial policies, but this item was not scored as it is not contained in the WHO Joint Statement. Two funders (Bill and Melinda Gates Foundation and the Department of Veterans Affairs – Office of Research and Development) were assessed during the pilot phase. No changes to the assessment tool and rating guide were required after the pilot’s completion. The pilot phase data was later integrated into the results. This study was prospectively registered with Open Science Framework (DOI <u>10.17605/OSF.IO/S8PTB</u>); the protocol, including all assessment tools, guides, and funder correspondence are available on GitHub.[27]

### Cohort selection

We compiled a list of large (>US$50 million annual spend) US medical research funders using data that was published in a peer-reviewed journal in 2016[11] and has been used for cohort selection in at least three separate studies of clinical trial policy transparency.[19–21] The list includes only noncommercial funders, categorized as either public or philanthropic.

Funders partially or wholly geographically located outside the United States as well as multilateral organizations such as WHO were excluded. Five public funders were excluded as they conducted human subject research but not clinical trials (National Aeronautics and Space Administration, Environmental Protection Agency); provided support for or regulation of clinical trials but did not fund them (National Science Foundation, Centers for Medicare and Medicaid Services); or engaged in pre-clinical testing but not clinical research (Department of Energy). Sixteen funders (7 public and 9 philanthropic) were initially identified as meeting the above inclusion criteria (>US$50 million annual spend, located in the US, and funding clinical trials).

Two additional medical research funder lists (one public and one philanthropic) were searched to identify any omitted funders. Forbes publishes an annual list of the largest US charities, ranked by donations received. The 2020 edition[28] was searched to identify omitted philanthropic funders of medical research. We filtered the Forbes list by category (Health), but no new funders were identified in the list’s entirety. For public funders, the 2018 “U.S. Investments in Medical and Health Research and Development” report was searched.[29] Again, no new funders were identified.

As the financial data from the initial list of 16 funders was a decade old (2013), we chose to manually update each funder’s expenditure estimate. Public funder expenditure amounts were brought current by searching the 2021 and 2022 congressional budgets, narrowing to “research” or “grants” where reported. For philanthropic funders, the most recent tax return Form 990 was used, which allowed grant spending to be isolated from other categories such as staff salaries and fundraising. Thus, some philanthropic funders’ actual research spending dropped below the US$50 million spending threshold, but they were retained in the cohort.

After cohort selection and during the assessment phase, the American Kidney Fund was found to primarily provide financial assistance for renal patients but does not sponsor clinical trials. They were excluded from the list. Additionally, after the assessment phase, the American Cancer Society were found to have no relevant clinical trial policies. Though they are listed under grant support for several trials[30, 31], their press office confirmed that they do not conduct clinical trials and do not serve as clinical trial sponsor. Thus, the resultant data from the American Cancer Society’s assessment was removed and they were excluded from the study, bringing the total number of funders to 14 (see Table 1).

**Table 1:** Funder list with updated expenditure Shading denotes a public funder

|  | Funder name | Acronym | Source | Expenditure (USD) |
| --- | --- | --- | --- | --- |
| 1 | National Institutes of Health | NIH | 2021 budget: https://officeofbudget.od.nih.gov/pdfs/FY22/cy/NIH%20Operating%20Plan%20-%20FY22%20Web%20Version.pdf | \$41.4 billion |
| 2 | Department of Defense (via Congressionally Directed Medical Research Programs) | DoD (CDMRP) | 2021 funding: https://cdmrp.army.mil/about/fundinghistory | \$1.8 billion |
| 3 | Bill and Melinda Gates Foundation | BMGF | 2020 annual report: https://docs.gatesfoundation.org/documents/2020_Annual_Report.pdf | \$1.79 billion |
| 4 | US Department of Veterans Affairs (Office of Research and Development) | VA (ORD) | 2021 budget: https://www.va.gov/budget/products.asp volume II, pg. 569 | \$795 million |
| 5 | Centers for Disease Control and Prevention | CDC | 2021 grant spending: https://taggs.hhs.gov/ReportsGrants/GrantsByActivityType | \$654 million |
| 6 | Agency for Healthcare Research and Quality | AHRQ | 2022 budget: https://www.ahrq.gov/sites/default/files/wysiwyg/cpi/about/mission/budget/2022/FY2022_CJ.pdf | \$488 million |
| 7 | Patient-Centered Outcomes Research Institute | PCORI | 2020 annual report: https://www.pcori.org/sites/default/files/PCORI-Annual-Report-2020.pdf | \$290 million |
| 8 | US Food and Drug Administration | FDA | 2021 grant spending: https://taggs.hhs.gov/ReportsGrants/GrantsByActivityType | \$173 million |
| 9 | American Heart Association | AHA | 2020 Form 990: https://www.heart.org/-/media/Files/Finance/20202021-IRS-Form-990-PDF.pdf | \$165 million |
| 10 | Leukemia and Lymphoma Society | LLS | 2020 Form 990: https://www.lls.org/sites/default/files/2022-02/FY21_LLS_990.pdf | \$153 million |
| 11 | Michael J. Fox Foundation for Parkinson's Research | MJFF | 2019 Form 990: https://www.michaeljfox.org/sites/default/files/media/document/2019-12_MJFF_FED_990_PUBLIC_DISCLOSURE_COPY_FOR_WEBSITE.pdf | \$97 million |
| 12 | Alzheimer's Association | AA | 2020 Form 990: https://www.alz.org/media/Documents/form-990-fy-2021.pdf | \$66 million |
| 13 | Juvenile Diabetes Research Foundation International | JDRF | 2020 Form 990: https://1x5o5mujiug388ttap1p8s17-wpengine.netdna-ssl.com/wp-content/uploads/2022/05/JDRF-990-FY21.pdf?_ga=2.50468649.1271124352.1657798787-187016285.1657020822 | \$28 million |
| 14 | Susan G. Komen Breast Cancer Foundation | SGK | 2020 Form 990: https://www.komen.org/wp-content/uploads/fy20-form-990-group.pdf | \$8 million |
| | | | | **Total = \$47 billion** |

The STROBE checklist for cross-sectional studies was used for study design and reporting to ensure all necessary elements were included.[32] No ethics approval was required by The London School of Economics as only publicly available institutional data were used.

### Rating

Two researchers per funding body independently searched funder websites and policy documents in June 2022. They each filled out the assessment tool using the rating guide, capturing relevant policy statements and hyperlinks. Scoring was binary (yes/no), and funders received no points for non-binding policies or those that did not cover all trials. Non-binding and partial coverage scores were noted in a separate scoresheet and appear in Table 2. Divergent ratings due to a rater’s oversight of a relevant policy element were reviewed by the team leader who determined the final score. Inter-rater reliability was not assessed as the aim was to capture all relevant policy statements.

**Table 2:** Full assessment results (includes CONSORT)

| Funder<br>(non-responders) | Pre-register | Updates | R-Results | Protocol | Journal pub | Trial ID | OA pub | Sanctions | Monitor reg. | Monitor res. | Pub report | CONSORT |
| --- | --- | --- | --- | --- | --- | --- | --- | --- | --- | --- | --- | --- |
| NIH | YES | YES | YES | YES | YES | NO | YES | YES | YES | YES | YES | no |
| FDA | YES | YES | YES | YES | S | S | YES | YES | S | S | NO | no |
| SGK | YES | YES | YES | NO | NO | NO | YES | YES | NO | NO | NO | no |
| CDMRP (DoD) | YES | NO | NO | NO | NO | NO | YES | NO | NO | NO | NO | no |
| BMGF* | S | NO | NO | NO | S | S | YES | NO | NO | NO | NO | journal |
| AHRQ | S | NO | S | NO | S | NO | YES | NO | NO | NO | NO | no |
| LLS | NO | NO | NO | NO | NO | NO | YES | NO | NO | NO | NO | no |
| CDC | NO | NO | NO | NO | NO | NO | YES | NO | NO | NO | NO | no |
| **Funder**<br>**(responders)** |  |  |  |  |  |  |  |  |  |  |  |  |
| VA (ORD) | YES | YES | YES | YES | NO | YES | YES | YES | YES | YES | NO | no |
| PCORI | YES | YES | YES | S | S | YES | YES | NO | YES | YES | YES | yes |
| MJFF | YES | NO | YES | NO | YES | NO | YES | S | NO | S | S | no |
| JDRF | YES | YES | S | NO | YES | S | YES | S | P | P | NO | no |
| AHA | YES | S | NO | S | YES | NO | YES | NO | NO | NO | NO | journal |
| AA | P | NO | NO | NO | YES | NO | YES | S | S | NO | NO | no |
| * denotes signatory of WHO Joint statement |  |  |  |  |  |  |  |  |  |  |  |  |
| S = funder supports/encourages practice without mandating it (non-binding) |  |  |  |  |  |  |  |  |  |  |  |  |
| P = funder policy item applies to a specific sub-set of trials (not all trials) |  |  |  |  |  |  |  |  |  |  |  |  |

Rater disagreements that were based on the same source text were referred to the adjudicator. Where applicable, we used precedents set in the Bruckner et al. 2022 adjudication document to settle ambiguous or difficult to score policy items. This ensured that our cohort is scored using the same criteria as were applied to the European funder cohort. All such decisions were recorded and added to the existing adjudication document. The final adjudication document, as well as all rounds of rating are available in GitHub.[27]

One frequently contended item was the timeframe for clinical trial registration. Several funders’ policies (BMGF, NIH, FDA) require registration within 21 days of trial initiation, which does not fulfill the “prospective” element in the WHO Joint Statement. However, the wording for this policy item in its entirety states the entry must be made “before the first subject receives the first medical intervention in the trial (*or as soon as possible afterwards*)”. Additionally, the FDAAA allows for trials to be registered up to 21 days after initiation. For this reason, all non-prospective trial registration policies that specified a 21-day window were still awarded the full point.

### Funder outreach

The press departments of all 14 funders were contacted by email with a copy of their completed score sheet, rating guide, and protocol. They were requested to flag possible errors and omissions. Each funder was contacted at least twice, two weeks apart. For the 6 funders who provided a response, the team leader corrected any errors or omissions where applicable. A third rater independently assessed the policies of all 8 non-responsive funders. Additionally, the assessments from the three largest non-responders (NIH, DoD, and BMGF) were compared against the raw data from the 2018 DeVito et al. study, whose evaluation also included these three funders.[19] This was a protocol deviation due to the low response rate amongst these large funders. Scores were updated for two items based on the NIH’s response[33] to the DeVito et al. team. Ratings for BMGF and DoD were consistent with the data from DeVito et al. study and original scores were retained.

Responses were received from 6/14 funders (43%) and ratings were adjusted for 18/66 items (6 funders x 11 items = 66 items total). In their responses, these six funders provided additional documents (award letters, grant terms and additional links to webpages) that contained policy items not found in public-facing documents. These were used to update the ratings for 18 items. The full scoresheet including changes made after funder response appears in the appendix. All responses and changes are also archived on GitHub.[27]

## Results

Our cross-sectional study demonstrated a large degree of variation among US funders’ adoption of WHO best practices. The most frequently adopted policy element was open access publication of research results, 14/14 (100%), followed by prospective trial registration, 9/14 (64%). Only two funders (14%) made public reports of grantee’s results reporting compliance, and only two funders (14%) required the trial ID to be included in all publications.

**Fig 1:**
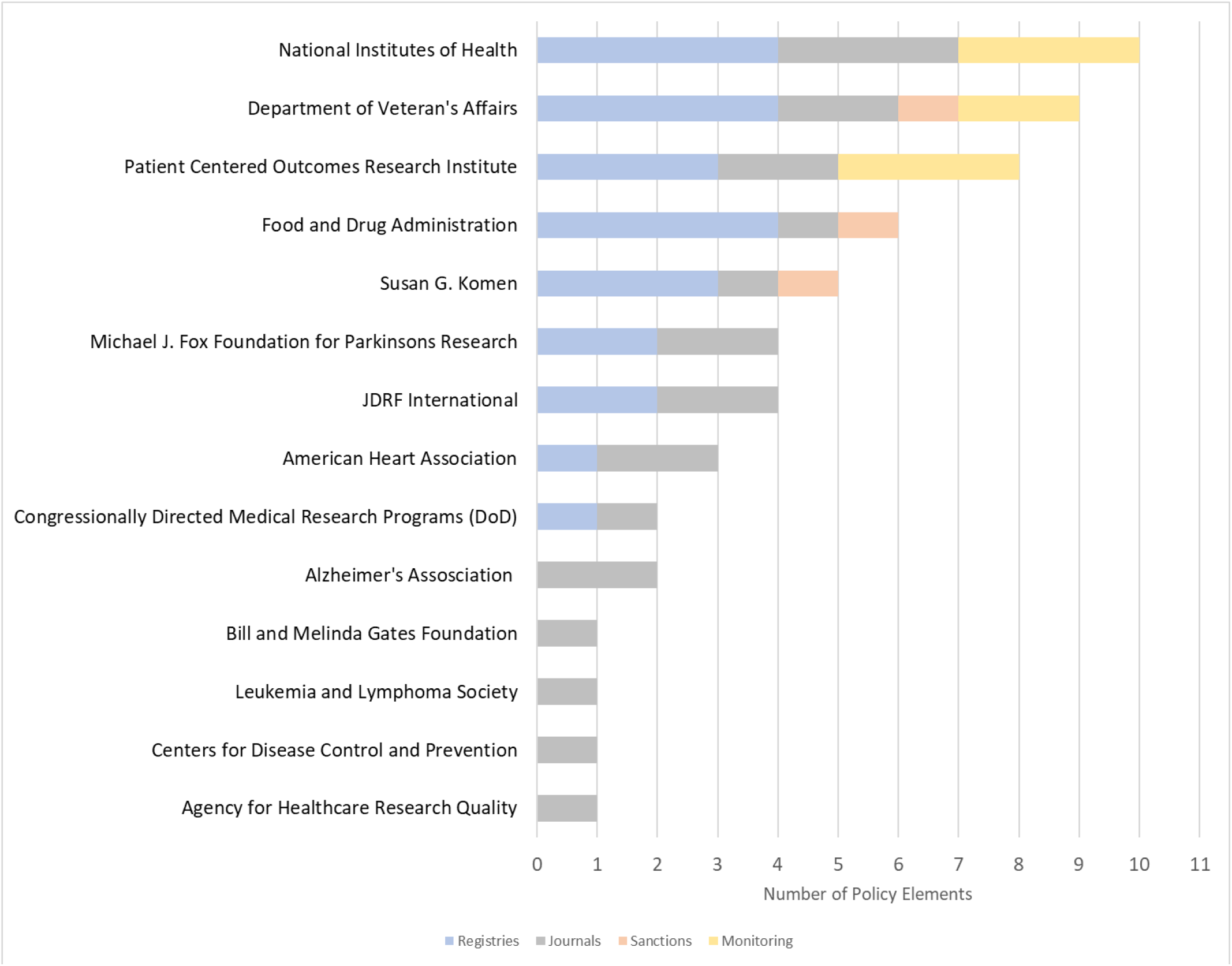
Number of policy elements adopted per funder

**Fig 2:**
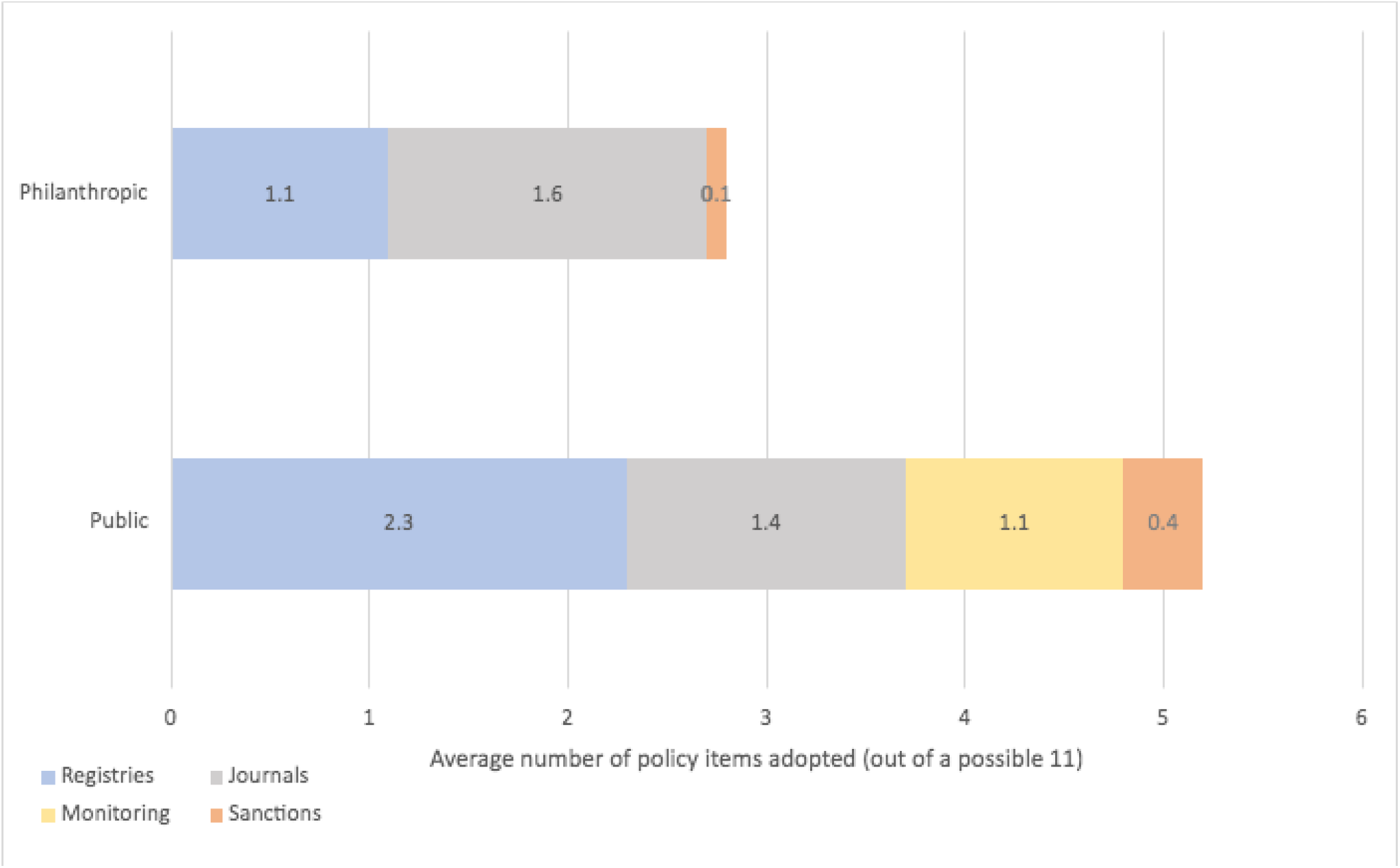
Public versus philanthropic performance by category

**Fig 3:**
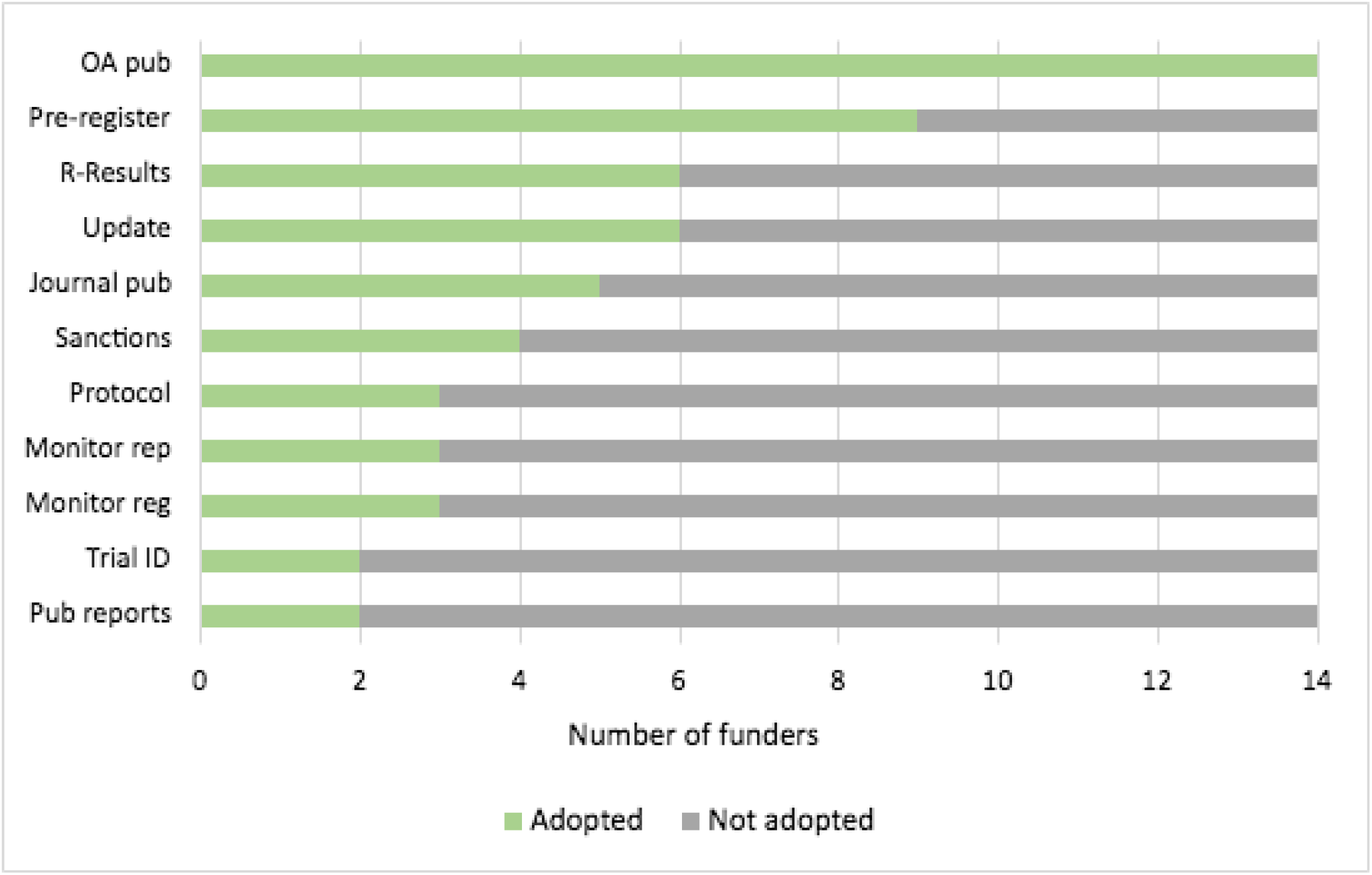
WHO best practice adoption, by number of funders adopting each practice

On average, the 14 largest US funders of medical research have adopted 4.1/11 (37%) of WHO best practices. Public funders adopted an average of 5.3/11 policy items (48%), while philanthropic funders averaged 2.8/11 (25%). The NIH had the greatest number of WHO best practices appearing in their policy documents, receiving a point for 10/11 items. Four funders (CDC, LLS, BMGF, AHRQ) had just one of the 11 WHO best practices: all required grantees to publish open access.

Compared with European and other global funders assessed in the 2023 O’Riordan et al. study[24], which found that 5/11 (45%) of WHO best practices were adopted on average amongst funders outside of the US, American institutions are faring slightly worse.

Just one funder referred to the Consolidated Standards of Reporting Trials (CONSORT) standards in their clinical trial results reporting policies.[25] Two additional funders refer to CONSORT in the publishing guidelines of their journals but did not require that their grantees publish in line with CONSORT.

## Discussion

Strong clinical trial registration and reporting policies, including the monitoring and public disclosure of these activities, can reduce medical research waste.[3] There is significant room for improvement among US funders of medical research, as on average, funders’ policies contain just 37% (4.1/11) of WHO best practices.

The most frequently adopted policy across US funders is open access publication (14/14). In 2013, the White House issued a memorandum directing federal agencies to develop open access policies for all federally funded research.[34] As a result, many federal agencies now have dissemination policies in place – all 7 federal funders had adopted with this particular WHO best practice. However, these policies are only as good as the funders’ expectation that grantees post and publish all results. Just 6/14 funders (43%) require that results are posted to ClinicalTrials.gov within 12 months of study completion, while only 5/14 (36%) funders require journal publication of research findings. One funder, Patient-Centered Outcomes Research Institute (PCORI) references CONSORT within its publication policies and had developed an exemplary guide to help grantees meet scientific integrity standards in their research reports.[35]

Public funders accounted for a significant portion of the medical research spending in this cohort: 95% of the US$47 billion, largely because of the NIH. Philanthropic funders, though accounting for half of funders assessed, spend far less research than public agencies. Thus, public funder policies hold greater weight, which this study did not adjust for.

Public funders’ policies were lacking in several areas. Very little information was contained within the FDA’s orphan products Request for Applications, and the CDC met only one WHO benchmark. The Department of Defense’s Congressionally Directed Medical Research Program (CDMRP) lacked several items, but it is possible that national security concerns stemming from military research might prevent full transparency. The NIH has a more developed and user-friendly grants section of their website, including a helpful compliance checklist and ACT decision tool. Though 10/11 items were identifiable in NIH policy documents, the section on clinical trial registration and reporting frequently linked to the entire 20-page FDAAA document[36] rather than itemizing each relevant policy. If full compliance is the goal, policies must be clear, concise, and contained directly on the NIH website.

Additionally, though on paper the NIH is the best performer in the cohort, in practice the NIH falls significantly short. In August 2022, the Office of Inspector General (OIG) released an audit of NIH-funded clinical trials’ compliance with federal reporting requirements.[37] The OIG audit of 72 studies found that just 15 extramural (21%) and 20 intramural (28%) trials had results submitted on time between 2019-2020. Though this is an improvement from the 8% on-time reporting rate in the 2015 Anderson et al. study, it is far from perfect. Despite comprehensive policies to the contrary, the report found: “NIH did not have adequate procedures for ensuring that responsible parties submitted the results of clinical trials, took limited enforcement action when there was noncompliance, and continued to fund new research of responsible parties that had not submitted the results of their completed clinical trials”. In response, NIH have pledged to improve procedures that will allow them to work with grantees on ClinicalTrials.gov registration and results submission compliance, as well as to reinforce their capacity to take corrective action.

Several funders signaled their intent to strengthen their policy during our outreach, and one funder, the Alzheimer’s Association, immediately changed their policy wording to better reflect WHO best practices.

Per the decisions in the adjudication document, these updated changes are not incorporated into the ratings, but all planned changes are noted in the supplementary material.

Clinical trial funders in the US have an opportunity to promote transparency, reduce research waste, and prevent publication bias through strong policies. Unfortunately, both public and philanthropic funders still fall short of WHO benchmarks.

### Limitations

We had a low funder response rate, 6/14 (43%). As many scores were adjusted after receiving responses and it is likely that some non-responding funders have more policy items than this team was able to locate publicly, our final ratings may not include all items. However, this underscores the value of having accessible, public-facing policies.

We received no response from 4 of the 5 largest funders. The NIH accounts for 86% of the US$47 billion in our cohort, and a response would provide validation for this significant funder. However, data from a previously published study[20] supported our findings for the 3 largest funders (NIH, CDMRP, and the Bill and Melinda Gates Foundation). Though the NIH was our top-performing funder, their failures to enforce their own policies are well documented, illustrating that there may be gaps between funder policies and funder practices, and highlighting the value of funders making audit reports public.

## Data Availability

All data produced are available online at Githb: https://github.com/emgamert/TransparencyUS2022

## References

[1] WHO. Joint statement on public disclosure of results from clinical trials, https://cdn.who.int/media/docs/default-source/clinical-trials/ictrp-jointstatement-2017.pdf (2017, accessed 29 July 2022).

[2] Ioannidis JPA. Clinical trials: what a waste. BMJ 2014; 349: g7089–g7089.

[3] Chalmers I, Glasziou P. Avoidable waste in the production and reporting of research evidence. The Lancet 2009; 374: 86–89.

[4] DeVito NJ, Goldacre B. Catalogue of bias: publication bias. BMJ Evid Based Med 2019; 24: 53–54.

[5] Turner EH, Cipriani A, Furukawa TA, et al. Selective publication of antidepressant trials and its influence on apparent efficacy: Updated comparisons and meta-analyses of newer versus older trials. PLoS Med 2022; 19: e1003886.

[6] Bruckner T. Clinical Trial Transparency: A Guide For Policy Makers, https://docs.wixstatic.com/ugd/01f35d_def0082121a648529220e1d56df4b50a.pdf (December 2017, accessed 29 July 2022).

[7] WMA Declaration of Helsinki - Ethical Principles for Medical Research Involving Human Subjects. Finland, https://www.wma.net/policies-post/wma-declaration-of-helsinki-ethical-principles-for-medical-research-involving-human-subjects/ (2013, accessed 29 July 2022).

[8] WHO. Strengthening clinical trials to provide high-quality evidence on health interventions and to improve research quality and coordination. In: Seventy-fifth World Health Assembly. Geneva, https://apps.who.int/gb/ebwha/pdf_files/WHA75/A75_ACONF9-en.pdf (2022, accessed 29 July 2022).

[9] National Institute of Allergy and Infectious Diseases. ClinRegs, https://clinregs.niaid.nih.gov/country/united-states#_top (2022, accessed 8 August 2022).

[10] U.S. National Library of Medicine. History, Policies, and Laws, https://clinicaltrials.gov/ct2/about-site/history (2021, accessed 9 August 2022).

[11] Viergever RF, Hendriks TCC. The 10 largest public and philanthropic funders of health research in the world: what they fund and how they distribute their funds. Health Res Policy Syst 2016; 14: 12.

[12] DHHS. 45 CFR 102.3. Title 45 Subtitle A, https://www.ecfr.gov/current/title-45/subtitle-A/subchapter-A/part-102/section-102.3 (2016, accessed 15 August 2022).

[13] FDA. ClinicalTrials.gov - Notices of Noncompliance and Civil Money Penalty Actions, https://www.fda.gov/science-research/fdas-role-clinicaltrialsgov-information/clinicaltrialsgov-notices-noncompliance-and-civil-money-penalty-actions (2022, accessed 15 August 2022).

[14] EBM DataLab. FDAAA Trials Tracker. University of Oxford.

[15] US National Library of Medicine. ClinicalTrials.gov FAQ, https://clinicaltrials.gov/ct2/manage-recs/faq (accessed 15 August 2022).

[16] Piller C. FDA and NIH let clinical trial sponsors keep results secret and break the law. Science, https://www.science.org/content/article/fda-and-nih-let-clinical-trial-sponsors-keep-results-secret-and-break-law (2020, accessed 24 August 2022).

[17] US Congress. Food and Drug Administration Amendments Act of 2007. 110–85, United States, https://www.govinfo.gov/content/pkg/PLAW-110publ85/pdf/PLAW-110publ85.pdf#page=95 (2007, accessed 15 August 2022).

[18] NIH. NIH Grants Policy Statement, https://grants.nih.gov/grants/policy/nihgps/nihgps.pdf (2021, accessed 15 August 2022).

[19] DeVito NJ, French L, Goldacre B. Noncommercial Funders’ Policies on Trial Registration, Access to Summary Results, and Individual Patient Data Availability. JAMA 2018; 319: 1721.

[20] Whitlock EP, Dunham KM, DiGioia K, et al. Noncommercial US Funders’ Policies on Trial Registration, Access to Summary Results, and Individual Patient Data Availability. JAMA Netw Open 2019; 2: e187498.

[21] Bruckner T, Rodgers F, Styrmisdóttir L, et al. Adoption of World Health Organization Best Practices in Clinical Trial Transparency Among European Medical Research Funder Policies. JAMA Netw Open; 5. Epub ahead of print 1 August 2022. DOI: 10.1001/jamanetworkopen.2022.22378.

[22] Prayle AP, Hurley MN, Smyth AR. Compliance with mandatory reporting of clinical trial results on ClinicalTrials.gov: cross sectional study. BMJ 2012; 344: d7373–d7373.

[23] Anderson ML, Chiswell K, Peterson ED, et al. Compliance with Results Reporting at ClinicalTrials.gov. New England Journal of Medicine 2015; 372: 1031–1039.

[24] O’Riordan M, Haslberger M, Cruz C, et al. Are European Clinical Trial Funders Policies on Clinical Trial Registration and Reporting Improving? – A Cross-Sectional Study. MedRxiv.

[25] Schulz KF, Altman DG, Moher D. CONSORT 2010 Statement: updated guidelines for reporting parallel group randomised trials. Trials 2010; 11: 32.

[26] CONSORT. Endorsers Journals and Organizations, http://www.consort-statement.org/about-consort/endorsers1 (2022, accessed 21 July 2022).

[27] Gamertsfelder E. GitHub, https://github.com/emgamert/TransparencyUS2022 (accessed 9 August 2022).

[28] Forbes. The 100 Largest U.S. Charities, https://www.forbes.com/lists/top-charities/ (2021, accessed 17 July 2022).

[29] ResearchAmerica. U.S. Investments in Medical and Health Research and Development: 2013-2017, https://www.researchamerica.org/sites/default/files/Policy_Advocacy/2013-2017InvestmentReportFall2018.pdf (2018, accessed 17 July 2022).

[30] D’Souza A, Szabo A, Flynn KE, et al. Adjuvant doxycycline to enhance anti-amyloid effects: Results from the dual phase 2 trial. EClinicalMedicine 2020; 23: 100361.

[31] Tang M, Chen M, Bruera E, et al. Association among rescue neuroleptic use, agitation, and perceived comfort: secondary analysis of a randomized clinical trial on agitated delirium. Supportive Care in Cancer 2021; 29: 7887–7894.

[32] von Elm E, Altman DG, Egger M, et al. The Strengthening the Reporting of Observational Studies in Epidemiology (STROBE) Statement: Guidelines for reporting observational studies. International Journal of Surgery 2014; 12: 1495–1499.

[33] DeVito et al 2018 -Non-Commercial Funder Policy Audit Archive, https://figshare.com/s/276f9dc0b37d8ecd0ab0 (2018, accessed 16 August 2022).

[34] Holden JP. Memorandum for the Heads of Executive Departments and Agencies: Increasing Access to the Results of Federally Funded Scientific Research. United States, https://rosap.ntl.bts.gov/view/dot/34953 (2013, accessed 15 August 2022).

[35] PCORI. Draft Final Research Report: Instructions for Awardee, https://www.pcori.org/sites/default/files/PCORI-Draft-Final-Research-Report-Instructions.pdf (2021, accessed 13 August 2022).

[36] US National Library of Medicine. FDAAA 801 and the Final Rule, https://clinicaltrials.gov/ct2/manage-recs/fdaaa (2022, accessed 8 August 2022).

[37] Office of Inspector General. The National Institutes of Health Did Not Ensure That All Clinical Trial Results Were Reported in Accordance With Federal Requirements, https://oig.hhs.gov/oas/reports/region6/62107000.asp (mAugust 2022, accessed 16 August 2022).

